# Very high frequencies of Familial Breast Cancer, low BRCA1/2 mutations and high Luminal molecular subtypes in Eastern Saudi Arabia necessitate the hunt for novel breast cancer hub genes

**DOI:** 10.1101/2023.10.03.23296209

**Authors:** Tahani Al-Qurashi, Sarah Balghonaim, Haya Albunyan, Hattan Alduhailan, Latiffah Almqhawi, Irtaza Fatima Zafar, Rehab Abubaker, Anees R Malik, Hani Mustafa, Nawaf Alanazi, Zafar Iqbal

## Abstract

This study was carried out to find out incidence of familial breast cancer, frequencies of different histological and molecular subtypes of breast cancer, and proportion of breast cancer patient with BRCA1/2 mutations, to plan for its implementation in early cancer screening and personalized treatment in Eastern Saudi Arabia. The breast cancer incidence increased from 2009-2016 and peaked in 2012. Ductal carcinoma (80%) was the most common histological type and Luminal A/B the most common (74%) molecular types among all patients. The familial and non-familial breast cancer groups were compared based on their demographic and clinical characteristics. The mean age at diagnosis was 51.6±13.5 and 53.5±12.7 for familial and non-familial breast cancer patients respectively. A total of 10 (24.3%) of familial breast cancer patients were tested for BRCA gene mutations, and 5 (12.2%) were abnormal, making it 4.3% of total patients. In the familial group, significant correlation (p 0.04) was found between patients with age less than 50 and with more than one family member affected with breast or ovarian cancer. Significance was found between unilateral familial breast cancer and bone metastasis (p 0.004). In both familial and non-familial groups, significant correlation was found between unilateral breast cancer and liver metastasis (0.005 and 0.017 respectively). There was a significant association between unilateral non-familial breast cancer and luminal A molecular type (p 0.012). Analysis of TNM staging showed that (61%) of familial breast cancer patients were diagnosed at stage II. In conclusion, familial breast cancer incidence is increasingly high in Al-Ahsa region as compared to western populations which could be due to consanguineous marriage and social behaviors. It warrants further studies to explore familial breast cancer hub genes in Eastern Saudi Arabia and its implementation in early diagnosis and patient-tailored treatment of breast cancer.

## 1. Introduction

Breast cancer is one of the most common cancers in women around the world ^1^. It is the second leading cause of cancer death in women aged 40 to 49, responsible almost one third of deaths before 50 years of the age^2^. There are many risk factors of breast cancer some of which are uncontrollable (age, family history, genetic mutations etc.) while others are controllable (for example, alcohol consumption, smoking, obesity and poor physical activity)^3^. Patient awareness, counselling of affected families, effective screening, early diagnosis and patient-tailored treatment can help not merely in reducing these risks but also help in effective treatment and even provide cure in many patients^1-3^. This is because some molecular subtypes of breast cancer like Luminal A have very good prognosis, excellent response to anti-cancer therapy, low probability of metastasis as well as relapses ^4^ while some other subtypes like Triple-negative breast cancer (TNBC) have poor prognosis, non-satisfactory response to chemotherapy and high frequencies of metastasis as well as relapses ^5^.

Genetic predisposition to breast/ovarian cancer and positive family history are of the most important risk factors for breast cancer. Globally, almost 5-10% cases in breast cancer patients have positive family history ^6^. This group of inherited or familial breast cancer carries mutations in several breast cancer susceptibility genes including BRCA1, BRCA2, TP53, CDH1, PTEN, STK11, NF1, ATM, CHEK2, PALB2, BARD1, RAD51C and RAD51D, as recommended by recent National Comprehensive Cancer Network USA (NCCN) guidelines^7^. However, high-penetrance genes like BRCA1 and BRCA2 cover only 5-10% of inherited breast cancers while majority of familial breast cancer cases are due to low penetrance genes that are inherited in autosomal recessive fashion^8^. In populations with high frequencies of consanguinity, there are higher chances convergence of recessive mutations and subsequently inherited diseases ^9^. A very recent study have reported high incidence of familial breast cancer in Saudi Arabia and other Arabian Gulf countries due to significantly high rates of consanguineous marriages ^9^.

There are no studies available about frequencies inherited breast cancer, BRCA1/2 mutations and its molecular subtypes in Eastern region and utilization of the resulting data in early screening, treatment, prevention and clinical management although such studies have been reported from other regions of Saudi Arabia^10^. Therefore, objective of this was to find out incidence of familial breast cancer in Al-Ahsa area of the Eastern region, frequencies of causative BRCA1/2 mutations, and estimate the proportion of familial breast cancer patients that might have recessive BRCAx (non-BRCA1/2 genes) in order to plan new screening strategies for early detection of breast cancer in high-risk Eastern region of Saudi Arabia.. Furthermore, frequencies of prognostically important common histological and molecular subtypes of breast cancer as well as their clinical characteristics were also investigated to see how early diagnosis of breast cancer by employing molecular biological methods can help in devising strategies for patient-tailored treatment of breast cancer patients in Al-Ahsa and other parts of Eastern region in Saudi Arabia.

## 2. Methods

The study included aall breast cancer patients who attended the breast oncology clinic at King Abdulaziz Hospital (KAH), Al-Ahsa, Saudi Arabia from 2007-2022. It was carried out at King Abdulaziz Hospital and King Saud Bin Abdulaziz University of Health Sciences, Al-Ahsa, Saudi Arabia. The study was approved by Scientific Committee and Institutional Review Board of participating centres.

Patients’ files and electronic record data were thoroughly examined, and all data related to clinical testing, treatment response and demographics was documented in the patient data form.

For detecting BRCA1/2 mutations, all procedures were adopted as described earlier ^11^. Briefly, 5 ml of peripheral blood was collected from the patients in EDTA tubes (BD Vacutainer Systems, Franklin Lakes, NJ, USA). DNA was extracted using QIAamp DNA Blood Mini Kit (QIAGEN, Germany. DNA was quantified by utilizing NanoDrop Spectrophotometer (NanoDrop Technologies, Inc., Wilmington, DE, USA) and aliquots of 70–80 ng/μL of DNA prepared for BRCA1/2 mutation detection by next generation sequencing (NGS). BRCA1/2 genes were screened by next-generation sequencing on Illumina NextSeq500 instrument (Illumina, San Diego, CA, USA) by employing manufacturer’s protocol. The variants screened through NGS were confirmed by Sanger sequencing using ABI 3730xl Analyzer (Applied Biosystems/Life Technologies, Carlsbad, CA, USA).

All data from King Abdulaziz Hospital’s information system BESTCare 2.0 system (ezCaretech, Seoul, South Korea) was retrieved and analyzed using IBM SPSS Statistics (Version 27).

## 3. Results

This study included 115 patients diagnosed with breast cancer in King Abdul-Aziz Hospital in Al-Ahsa Saudi Arabia. The breast cancer incidence increased from 2009-2017 and peaked in 2012 with 19 cases followed by 17 in 2016 (17.7%). Low number of patients from 1990 to 2008 is attributed to limited access to data from those years.

The 115 patients were further divided based on gender into females and males with a frequency and percentage of 113 (98.3%) and 2 (1.7%) respectively. 60 (53.1%) of the patients were diagnosed at ages above 50 years (table 1)

**Table 1.**
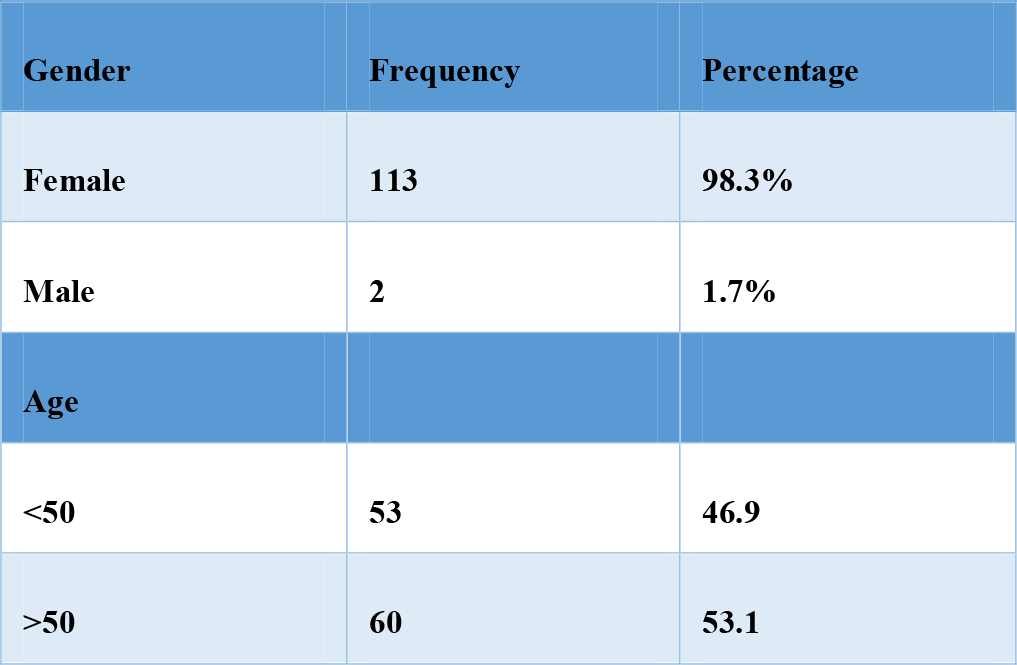
Distribution of Breast Cancer Patients According to Gender and Age.

| Gender | Frequency | Percentage |
| --- | --- | --- |
| Female | 113 | 98.3% |
| Male | 2 | 1.7% |
| Age |  |  |
| <50 | 53 | 46.9 |
| >50 | 60 | 53.1 |

Patients were divided and compared based on family history of breast/ovarian cancer into familial (group 1), non-familial (group 2) and patients whose family history data were not available (group 3) as shown in Table 2 (Figure 1). Family history data were available for 78 (67.8%) patients. 41 (36%) patients had a family history of breast or ovarian cancer while 37 (32%) patients did not. 37 (32%) patients had no family history data.

**Table 2.** Grouping of Breast Cancer Patients Based on Family History.

|  | Familial<br>(Group 1) | Non-Familial<br>(Group 2) | No Family<br>History Data<br>(Group 3) |
| --- | --- | --- | --- |
| Percentage | 41 (36%) | 37 (32%) | 37 (32%) |

**Figure 1.**
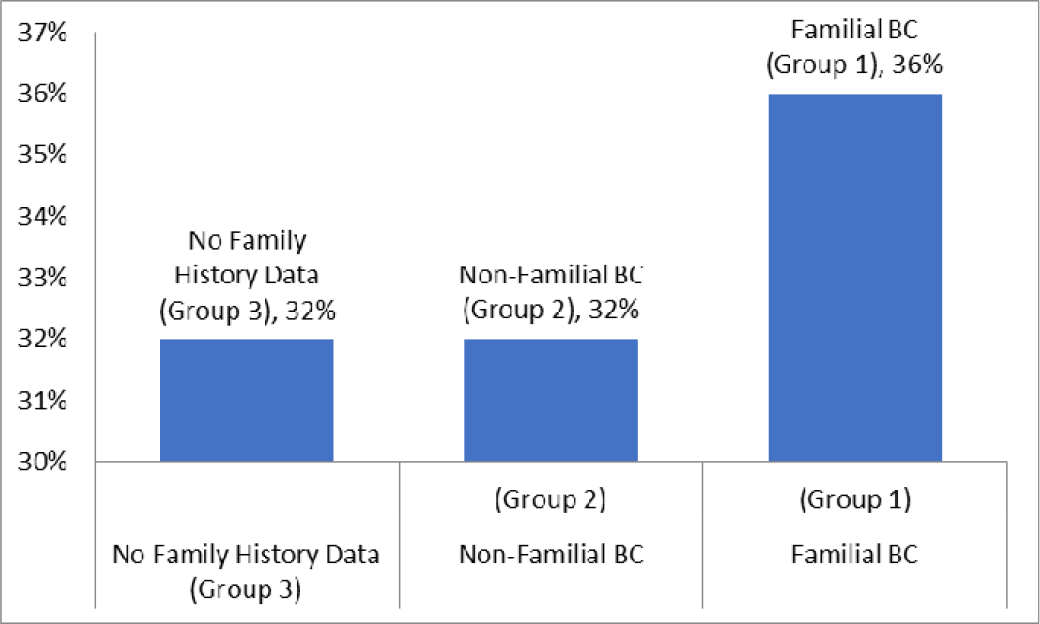
Grouping of Breast Cancer Patients Based on Family History

The mean age at diagnosis was 51.6 ±12.8 for all patients, 51.6±13.5 for familial breast cancer patients (group 1) and 53.5±12.7 for non-familial breast cancer patients (group 2) as shown in Table 3. Group 1 and 2 showed similar mean age at diagnosis (Figure 2).

**Table 3.** Age Distribution among different patient groups.

| Group | Mean | Standard Deviation |
| --- | --- | --- |
| Group 1 (familial) | 51.6 | 13.5 |
| Group 2 (Non-familial) | 53.5 | 12.7 |
| All groups | 51.6 | 12.8 |

**Figure 2.**
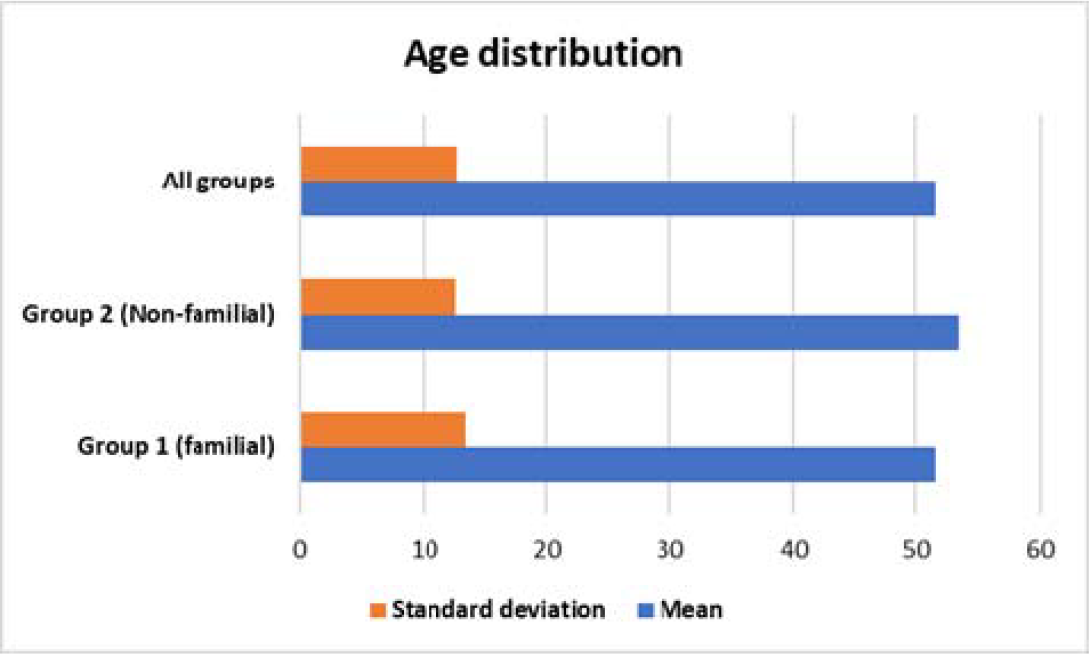
Age Distribution among different patient groups in breast cancer

Table 4 shows the subdivision of the familial breast cancer (patients (group1, 41 or 36% of the total breast cancer patients) based on the number of affected family members with ovarian or breast cancer into group 1A with only one family member affected (87.5%) and group 1B with more than one family member affected (12.5%).

**Table 4.**
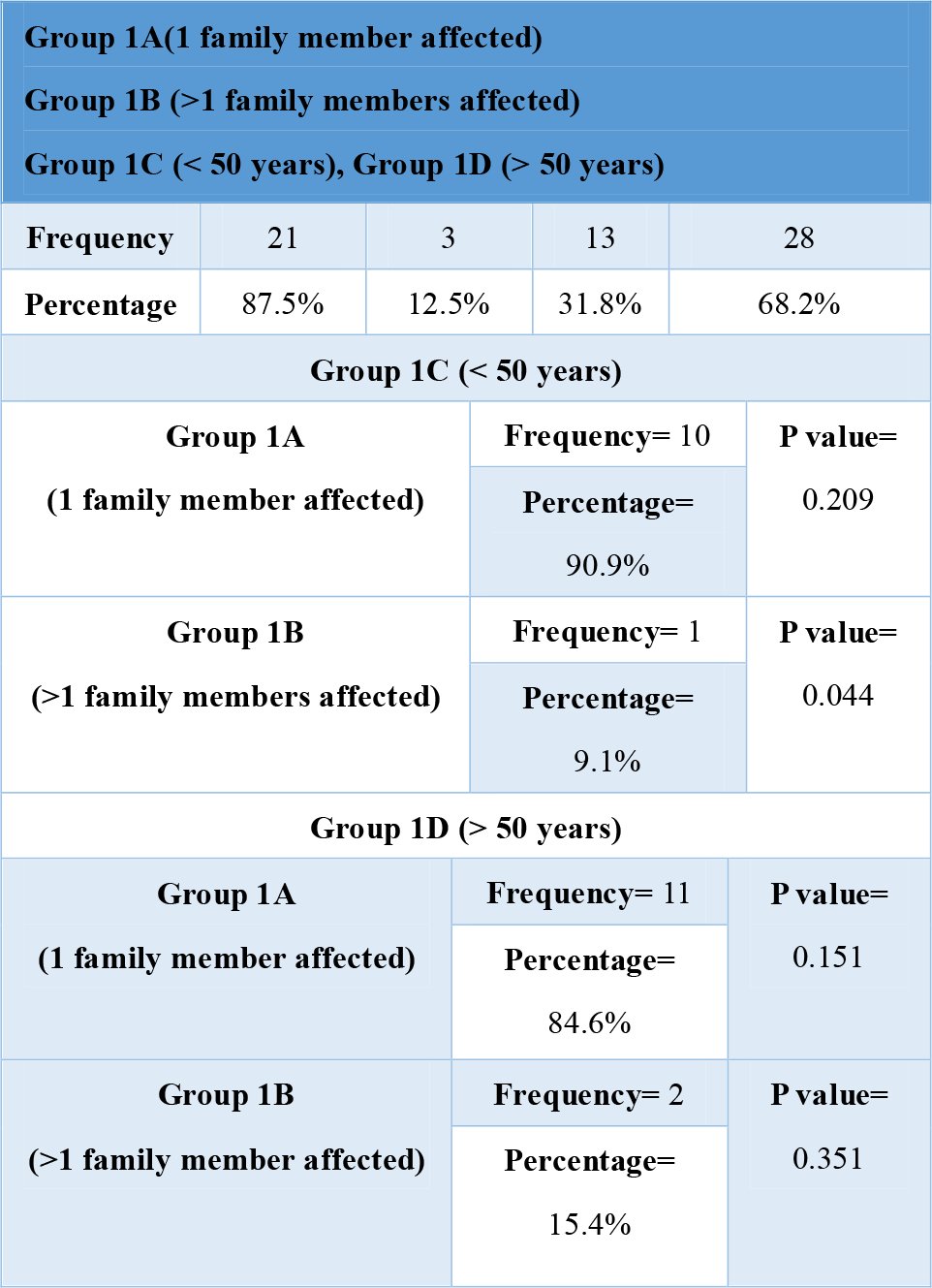
Division of Familial Breast Cancer Patients (Group 1) into Sub-groups Based on Number of Affected Family Members and Age.

We also correlated age of familial breast cancer patients with number of family members with breast/ovarian cancers. Whereas when basing on age, group 1 patients (familial breast cancer group, 41 or 36% of the total breast cancer patients) were further divided into group 1C with an age less than 50 years (31.8%) and group 1D with an age more than 50 years (68.2%). Only 10 (90.9%) patients were found to have 1 affected family member with age less than 50 years (group 1A and 1C). Only 1 (9.1%) patient had 2 affected family members with age less than 50 years (group 1B and 1C) with a statistically significant p-value of 0.044. Moreover, 11 (84.6%) patients had 1 affected family member and aged more than 50 years (group 1A and 1D). Only 2 (15.4%) patients had 2 affected family members and aged more than 50 years (group 1B and 1D). Therefore, percentage of familial breast cancer patients with 2 or more affected family members and age less than 50 years was very low (Table 7).

Overall, Luminal type A/B was most common (74%, two third) in our breast cancer patients (Figure 3). Luminal A molecular type was significantly correlated with breast cancer laterality of non-familial breast cancer patients with a p-value of 0.021 (Table 5).

**Table 5.** Significant Correlation between Breast Cancer Laterality and Luminal-A Molecular Type in Non-Familial Breast Cancer Patients.

| Non-Familial Breast Cancer Patients (Group 2) |  |
| --- | --- |
| Breast Cancer Laterality | P Value |
| Unilateral | 0.021 |

**Figure 3:**
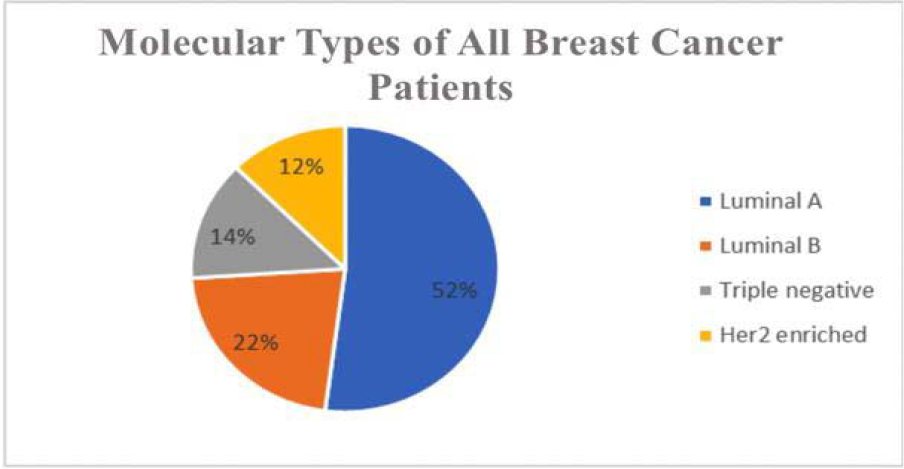
High frequencies of Luminal A/B types of Breast Cancer.

Table 6 demonstrates the significant correlation between breast cancer laterality and bone metastasis only in familial breast cancer patients with a p-value of 0.004. Similarly, liver metastasis was significantly correlated with breast cancer laterality with a p-value of 0.005 in familial breast cancer patients and 0.017 in non-familial breast cancer patients.

**Table 6.** Significant correlation Between Metastasis sites and Breast Cancer Laterality among Familial and Non-familial Breast Cancer Patients.

| Familial Breast Cancer Patients (Group 1) |  |  |
| --- | --- | --- |
| Breast Cancer Laterality | Bone metastasis | Liver metastasis |
| Unilateral | 0.004 | 0.005 |
| Non-familial Breast Cancer Patients (Group 2) |  |  |
| Unilateral | 0.120 | 0.017 |

**Table 7.** Breast Cancer Patients Tested for BRCA1/2 Gene Mutations.

|  | Number of patients tested for BRCA | Normal | Mutated |
| --- | --- | --- | --- |
| Group 1 (Familial) | 10 (24.3%) | 5 (12.3%) | 5 (12.3%) |
| Group 2 (Non-familial) | 4 (9.7%) | 4 (9.7%) | 0 |
| Group 3 (No family history data) | 7(17%) | 7 (17%) | 0 |

A total of 10 (24.3%) of group 1 (familial breast cancer) patients were tested for BRCA1/2 and 5 (12.2%) found mutated (4.3% of total breast cancer patients). On the other hand, none of the 4 (9.7%) patients of group 2and 7 (17%) from group 3 tested for BRCA1/2 were detected with BRCA1/2 mutations (Table 7).

Table 8 shows more details about the patients who were tested positive for BRCA1/2 in reference to the mutation type, mutation class, TNM staging, molecular type, histological type, age and number of affected family members. Mean age of patients with BRCA1/2 is much lower (38) than overall mean age (51.6), mean age for familial BC (51.6) and mean age for non-familial BC (53.5%). Ductal carcinoma was found in 3 cases, and mammary carcinoma was found in 2 cases. Two out of the patients had stage III breast cancer while the rest had stage II. The triple negative was found in 2 patients one with BRCA1 and one with BRCA2 mutations. Luminal A was found in 2 cases, and luminal B was found in only one case. Out of the 5 patients, two had a nonsense mutation, two had a substitution mutation and only one had a deletion mutation.

**Table 8.** Clinical Characteristics of Familial Breast Cancer Patients Who Tested Positive for BRCA1/BRCA2 Gene Mutation.

|  | Gene | Mutation Class | TNM | Mutation type | Molecular type | Histological type |
| --- | --- | --- | --- | --- | --- | --- |
| 1 | BRCA1 | Nonsense | III | c.454G>A p.(Trp1508) | Triple negative | Mammary |
| 2 | BRCA2 | Substitution | II | IVS9-17G>A | Luminal A | Ductal |
| 3 | BRCA2 | Substitution | II | S1123G (3595A>G) | Triple negative | Ductal |
| 4 | BRCA2 | Nonsense | II | c.9976A>T p.(Lys3326) | Luminal A | Ductal |
| 5 | BRCA2 | Deletion | III | c.2808_2811del ACAA | Luminal B | Mammary |

Among the five familial breast cancer patients (group 1) who had mutations in their BRCA gene, 4 (80%) patients had mutated BRCA2 genes and only 1 (20%) patient had a mutated BRCA1 gene as shown in Table 9.

**Table 9.** Frequency and Percentage of BRCA1 and BRCA2 among Patients Who Tested Positive for BRCA Gene Mutations.

| Gene | Frequency | Percentage |
| --- | --- | --- |
| BRCA1 | 1 | 20% |
| BRCA2 | 4 | 80% |

The most three frequent diseases in familial (group 1) and non-familial (group 2) breast cancer patients were listed in Table 10. It was noticed that the most three frequent diseases were diabetes mellites (41%) in group 1 and (41%) in group 2, hypertension (35%) in group 1 and (40%) in group 2, and bone diseases including osteoarthritis, osteomyelitis, osteoporosis and calcification (24%) in group 1 and (19%) in group 2.

**Table 10.**
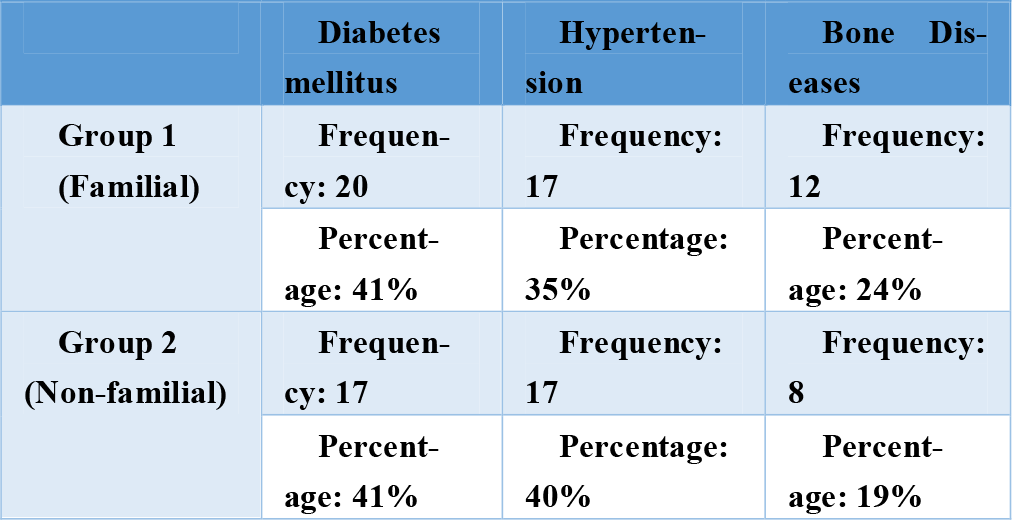
Most Frequent Related Disease in Familial and Non-Familial Breast Cancer Patients.

The most common metastasis sites in breast cancer patients were bone (32.7%) followed by liver (23%), lung (21.1%), brain (9.7%), the other breast (5.8%), lymph nodes (5.8%), and skin (1.9%). There was no significant difference among study groups when compared (Table 11 and Figure 12).

**Table 11.** Metastasis Sites of All Breast Cancer Patients.

| Table 11. Metastasis Sites of All Breast Cancer Patients |  |  |  |
| --- | --- | --- | --- |
|  | Site of Metastasis | Frequency | Total Percentage |
| 1 | Bone | Total= 17 | 32.7 |
|  |  | Group 1= 5 |  |
|  |  | Group 2= 5 |  |
|  |  | Group 3= 7 |  |
| 2 | Lung | Total= 11 | 21.1 |
|  |  | Group1= 3 |  |
|  |  | Group 2= 2 |  |
|  |  | Group 3= 6 |  |
| 3 | Lymph nodes | Total= 3 | 5.8 |
|  |  | Group1= 2 |  |
|  |  | Group 3= 1 |  |
| 4 | Liver | Total= 12 | 23 |
|  |  | Group1= 3 |  |
|  |  | Group 2= 4 |  |
|  |  | Group 3= 5 |  |
| 5 | Skin | Total= 1 | 1.9 |
|  |  | Group 3= 1 |  |
| 6 | Brain | Total= 5 | 9.7 |
|  |  | Group1= 1 |  |
|  |  | Group 2= 2 |  |
|  |  | Group 3= 2 |  |
| 7 | Another breast | Total= 3 | 5.8 |
|  |  | Group2= 2 |  |
|  |  | Group3= 1 |  |

**Table 12.**
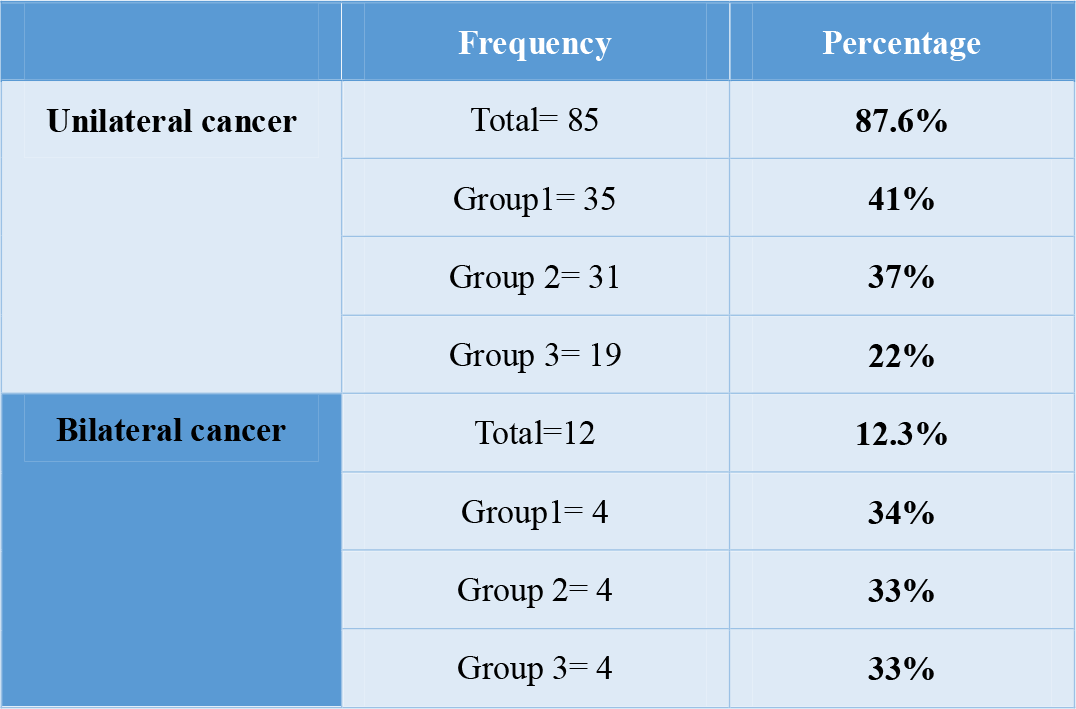
Distribution of Patients Based on Breast Cancer Laterality.

|  | Frequency | Percentage |
| --- | --- | --- |
| Unilateral cancer | Total= 85 | 87.6% |
|  | Group1= 35 | 41% |
|  | Group 2= 31 | 37% |
|  | Group 3= 19 | 22% |
| Bilateral cancer | Total=12 | 12.3% |
|  | Group1= 4 | 34% |
|  | Group 2= 4 | 33% |
|  | Group 3= 4 | 33% |

**Figure 11.1.**
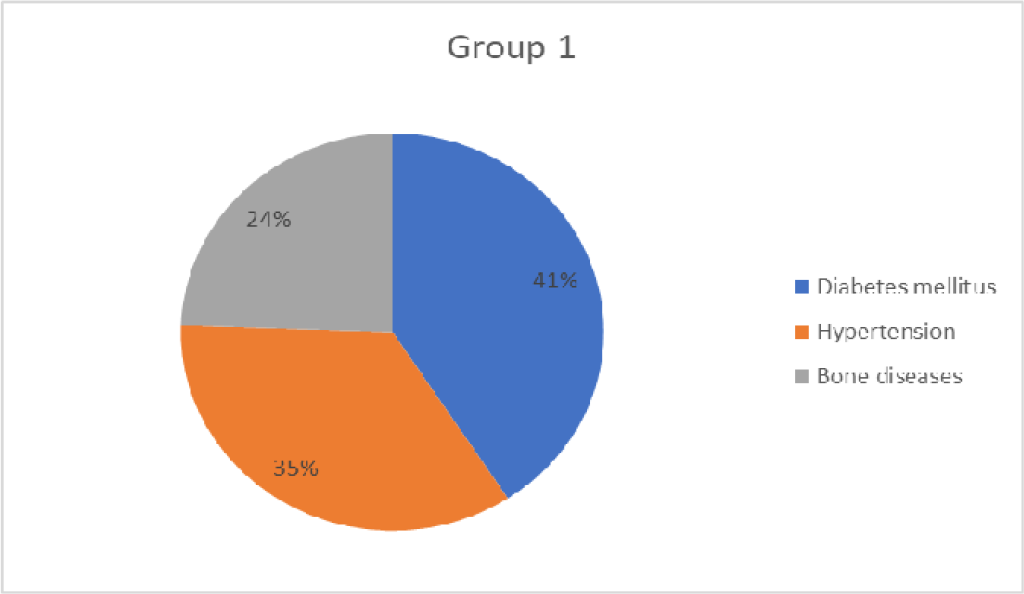
Most Frequent Related Disease in Familial Breast Cancer Patients

**Figure 11.2.**
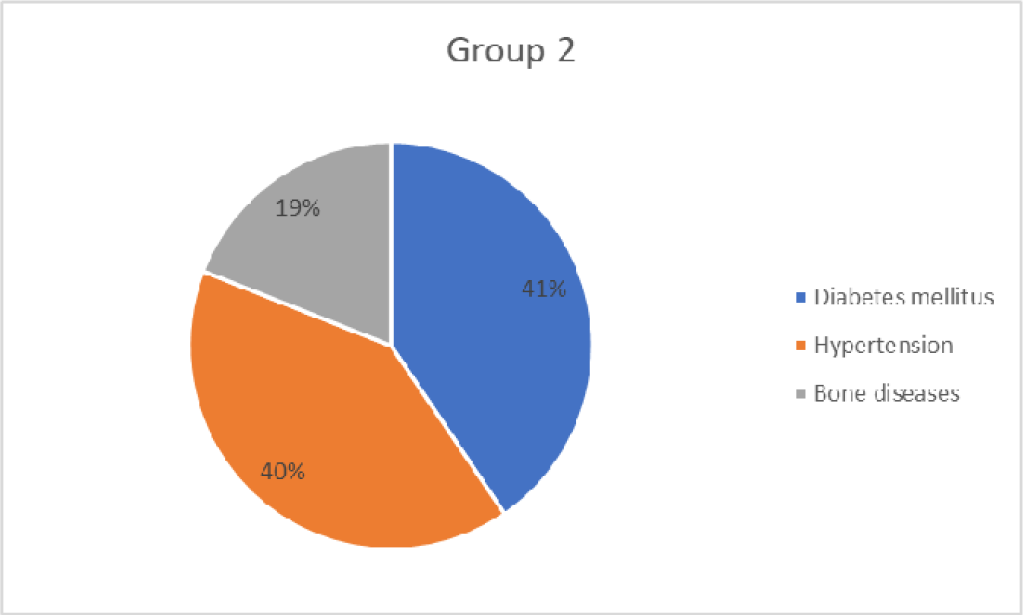
Most Frequent Related Disease in Familial Breast Cancer Patients

**Figure 12.**
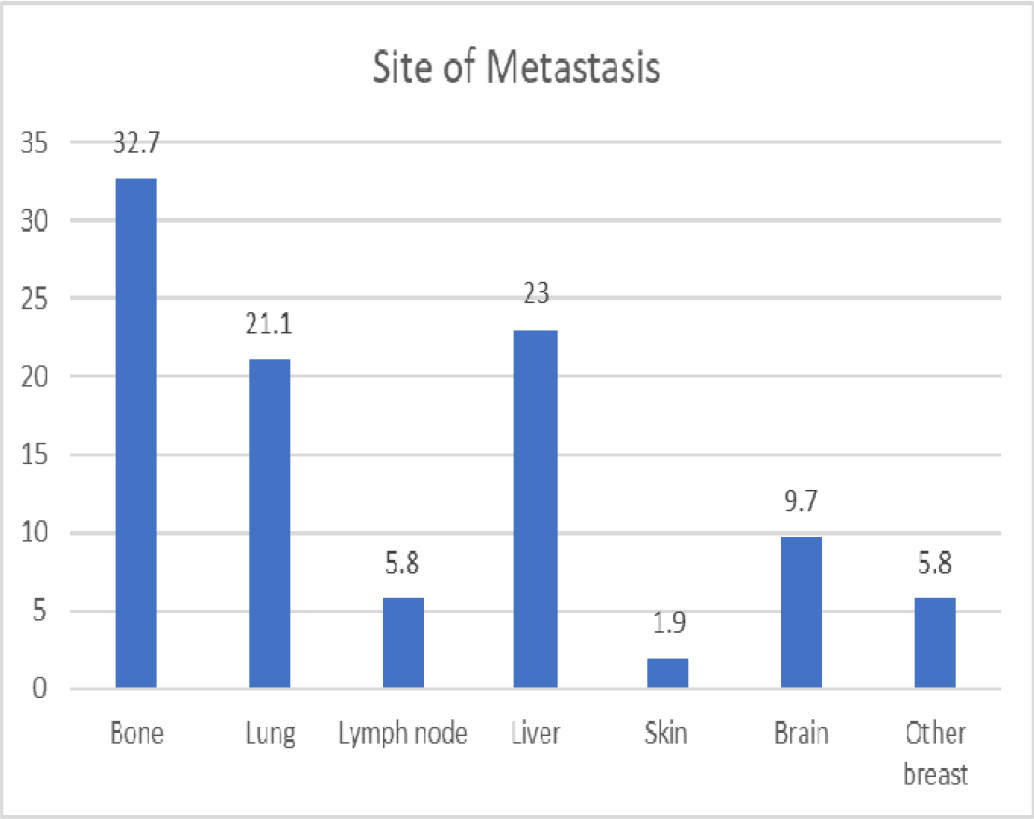
Percentage of Metastasis Sites of All Breast Cancer Patients

**Figure 13.1.**
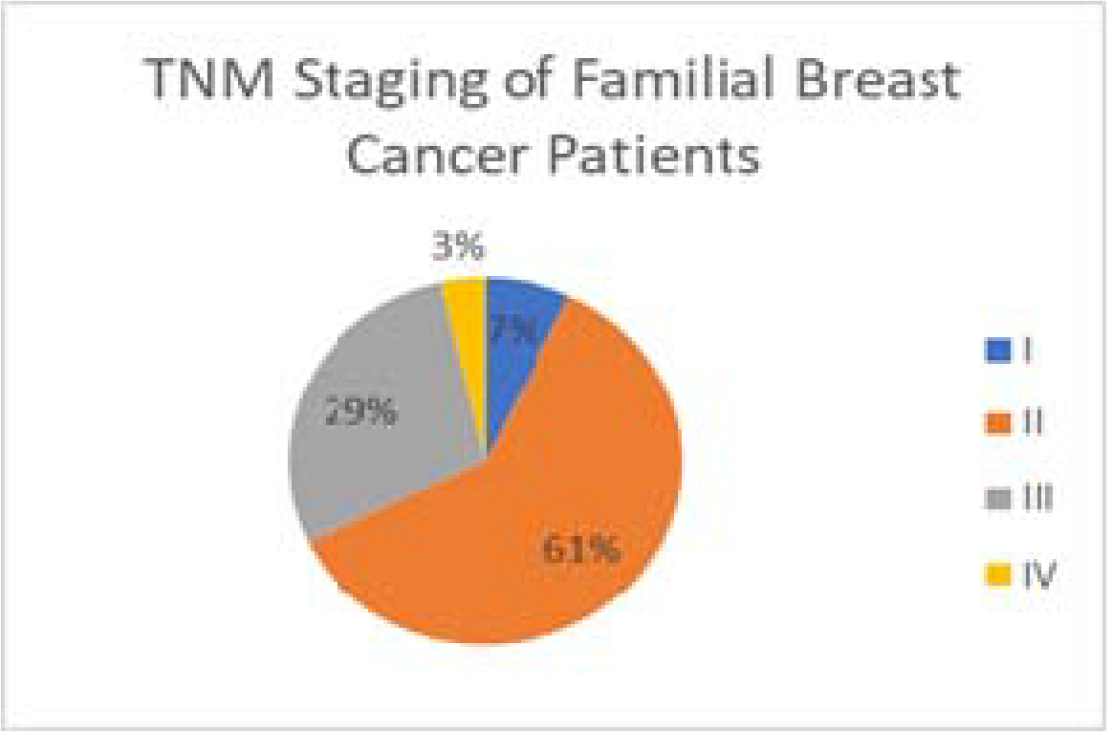
TNM Staging of Familial Breast Cancer Patients

**Figure 13.2.**
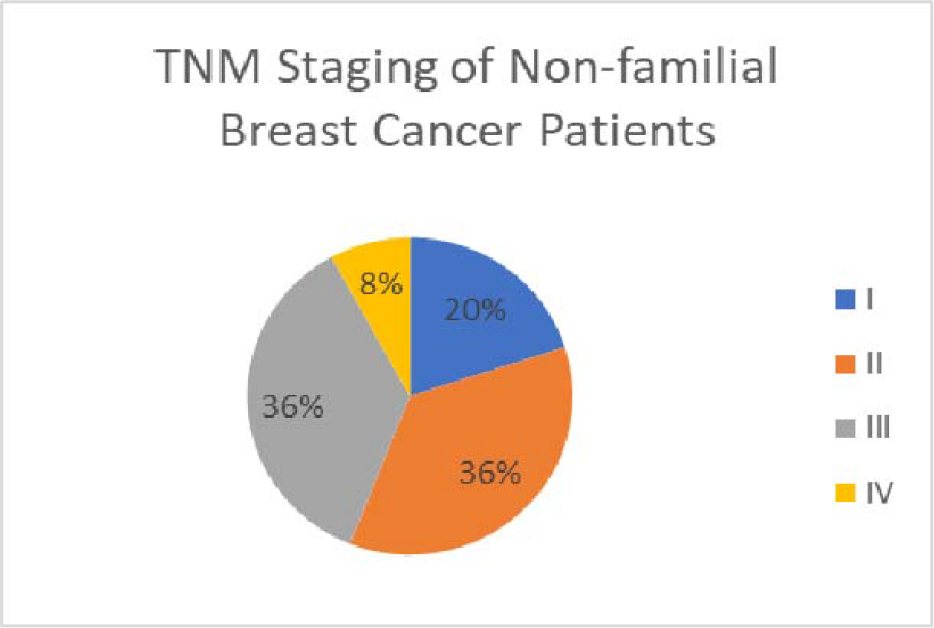
TNM Staging of Non-Familial Breast Cancer Patients

The majority (87.6%) of breast cancer patients had unilateral breast cancer while (12.3%) of the patients had bilateral breast cancer. No statistical significance was observed after comparing familial and non-familial breast cancer groups having unilateral or bilateral breast cancer (Table 13).

**Table 13.** TNM Staging of Familial and Non-familial Breast Cancer Patients.

| TNM staging | Total percentage |
| --- | --- |
| I | 13.2% |
| Familial= 2 |  |
| Non-familial= 5 |  |
| II | 49% |
| Familial= 17 |  |
| Non-familial= 9 |  |
| III | 32% |
| Familial= 8 |  |
| Non-familial= 9 |  |
| IV | 5.8% |
| Familial= 1 |  |
| Non-familial= 2 |  |

Table 13 shows the TNM staging of familial and non-familial breast cancer patients. The most common stage among both groups is stage II (49%) followed by stage III (32%), stage I (13.2%) and lastly stage IV (5.8%). Out of the familial breast cancer group, most patients had stage II (61%) then stage III (29%), stage I (7%) and stage IV (3%). Whereas in non-familial breast cancer group, stage II (36%) and III (36%) were equally higher than stage I (20%) and stage IV (8%).

Our research demonstrates a strikingly increased incidence of hereditary breast cancer in Eastern Saudi Arabia. Furthermore, the frequencies of BRCA1/2 gene mutations are lower as compared to other ethnic groups. Additionally, Luminal A/B breast cancer accounts for 74% of all breast cancer cases. This indicates the uniqueness of breast cancer patient population in Eastern parts of Saudi Arabia.

## 4. Discussion

This study was carried out on 115 breast cancer patients to compare familial and non-familial breast cancer and to find out the clinical characteristics of familial breast cancer patients. In this study, unilateral breast cancer was revealed to be the most common type found in 85 (73.9%) of the study population. However, this result does not match with a previous study suggesting that familial breast cancer was found to be bilateral.^11^

Out of 115 patients, 41 (36%) patients had a history of breast cancer in their families (group 1). This group was further subdivided into subgroups based on the number of family members affected with breast cancer and age. A significant correlation was found in a patient whose age was less than 50 years and had more than 1 family member affected (p-value of 0.04). Overall, these findings were in accordance with findings reported by Brandt et al ^12^ which suggested that individuals with several relatives affected with breast cancer have a probability to develop breast cancer at an early age. However, our study showed that the frequency of familial breast cancer was higher (36%) in women above 50 years of age. On the contrary, other studies had reported that a high percentage of familial breast cancer incidences were found in women less than 50 years of age.^10^ One of the reason for higher age of onset in our study could be the late diagnosis of familial breast cancer due to social reasons.

In this study, (36%) patients had familial breast cancer which was much higher than other ethnic groups of the world. In the UK, the incidence rate of females to develop familial breast cancer was between (6-19%).^13^ This could be due to high ratios of consanguineous marriages in KSA which has a frequency of (51.3%).^14^

Overall, 21 (18.2%) patients were tested for BRCA gene mutations. Out of the total breast cancer patients, only 5 (4.3%) were tested positive for BRCA1\2 mutations and all of these patients had positive family history for breast cancer. This frequency of BRCA1/2 mutations is lower than some other Asian populations ^15^ as well as other gulf countries, including some previous studies from other regions of Saudi Arabia ^10^. Nevertheless, our results are in accordance with investigations on Western populations with 5-10% frequency of BRCA1/2 mutations in breast cancer ^4^.

Out of the familial breast cancer group, (80%) of the positive cases have been reported with mutations in BRCA2, and (20%) with BRCA1. This result ties well with previous studies wherein the rate of mutation in BRCA 2 was higher in patients with familial breast cancer.^16^ In patients who had mutations in BRCA2, two of them had a substitution mutation, one had deletion mutation and two had a nonsense mutation. The patient who had mutated BRCA1 had a non-sense mutation. The total frequency of gene mutation in this study was low than reported in other studies.^15,16^ Similarly, mean age of patients with BRCA1/2 is much lower (38) than overall mean age (51.6) which is in accordance with previously reported data.

When comparing the metastasis site with cancer laterality, a significant correlation was found between bone metastasis and unilateral familial breast cancer (p-value 0.004). Similarly, bone metastasis was the most common metastasis site in breast cancer patients with (32.7%) of the population affected. Other studies carried out in Germany showed that (50-70%) of females with breast cancer developed bone metastasis during their disease course.^18^ Whereas when compared to the findings of other studies conducted on different cancer types in King Abdul-Aziz hospital at the same period of our study, lymphoma was associated more with spleen metastasis, lung cancer with a brain, and colorectal cancer with liver metastasis. In our studies, a significant association was found between liver metastasis and both familial and non-familial breast cancer patients (p-value of 0.005 and 0.017, respectively).

When analyzing the other diseases among breast cancer patients, diabetes mellitus, hypertension and bone diseases had been reported with the highest percentage. Diabetes mellitus affected (32%) of the total population, and this finding was in line with a previous study that had shown an increased risk of breast cancer among diabetic females.^19^ Furthermore, (29%) of the patients were found to be hypertensive in this study, and this goes along with the finding of a previous study that had shown that women with hypertension may have (15%) increased risk for breast cancer.^20^

Bone diseases such as osteoarthritis, osteomyelitis, osteoporosis, and calcification are the third most related diseases with breast cancer affecting (22%) of the study population. In the non-familial breast cancer patients, a significant correlation was found between unilateral breast cancer and the luminal A subtype (p-value 0.012). In contrast, a study carried out in Guangdong, China compared the molecular type and laterality of breast cancer had shown that luminal B (Her-2+) was significantly correlated to unilateral breast cancer.^21^

Analysis of TNM staging showed that (61%) of familial breast cancer patients were diagnosed at stage II. This could be due to either early diagnosis or slow progression of the disease, as supported by higher frequencies of Luminal A/B molecular subtypes of the breast cancer in our region, which is higher than other regions of Saudi Arabia and even from other ethnic groups in the world ^6-7, 10-13^. Very recently, Luminal type breast cancer patients have been found to have prognostically important novel PIK3CA mutations and low PTEN expression (in one third and one fourth patients, respectively) ^22^. In that study, PIK3CA mutations were associated with resistance to neoadjuvant chemotherapy but do not affect the response to neoadjuvant endocrine therapy^22-23^.

These findings indicate that additional research is necessary to investigate the familial breast cancer hub genes in Eastern Saudi Arabia and their potential application in the early detection and personalized treatment of breast cancer.

## 5. Conclusions

Our study shows a much higher frequency of familial breast cancer as compared to global frequencies of breast cancer patients with positive family history. As BRCA1/2 gene mutations frequencies are lower than other ethnic groups, there is very strong possibility of involvement of recessive mutant genes in familial breast cancers patients of Al-Ahsa region due to very high ratios of consanguineous marriages in KSA. This, along with overall 74% frequency of prognostically favourable Luminal A/B type breast cancer and high frequency of family history positivity indicates hunt for recessive breast-cancer susceptibility genes, their implication in early molecular diagnosis an timely treatment of specifically help Luminal A/B type family history positive breast cancer patients. Therefore, further studies involving next-generation pathology techniques to explore genetic basis of breast cancer in this area and their implication in patient-tailored treatment of breast cancer in high risk breast cancer population of Al-Ahsa is strongly recommended.

## Supporting information

Raw data of Breast Cancer Patients at KAH Al-Ahsa

## Data Availability

All data produced in the present work are contained in the manuscript

https://1drv.ms/f/s!Ai2Oaa5jGDDx1EBqRwtjhrkpEIGz?e=uB81Xk

